# Distinct genome-wide DNA methylation and gene expression signatures in classical monocytes from African American patients with systemic sclerosis

**DOI:** 10.1101/2022.03.17.22272588

**Authors:** Peter C. Allen, Sarah Smith, Robert C. Wilson, Jena R. Wirth, Nathan H. Wilson, DeAnna Baker Frost, Jonathan Flume, Gary S. Gilkeson, Melissa A. Cunningham, Carl D. Langefeld, Devin M. Absher, Paula S. Ramos

## Abstract

**Background:** Systemic sclerosis (SSc) is a multisystem autoimmune disorder that has an unclear etiology and disproportionately affects women and African Americans. Despite this, African Americans are dramatically underrepresented in SSc research. Additionally, monocytes show heightened activation in SSc and in African Americans relative to European Americans. In this study, we sought to investigate DNA methylation and gene expression patterns in classical monocytes in a health disparity population.

**Methods:** Classical monocytes (CD14++CD16-) were FACS-isolated from 34 self-reported African American women. Samples from 12 SSc patients and 12 healthy controls were hybridized on MethylationEPIC BeadChip array, while RNA-seq was performed on 16 SSc patients and 18 healthy controls. Analyses were computed to identify differentially methylated CpGs (DMCs), differentially expressed genes (DEGs), and CpGs associated with changes in gene expression (eQTM analysis).

**Results:** We observed modest DNA methylation and gene expression differences between cases and controls. The genes harboring the top DMCs, the top DEGs, as well as the top eQTM loci were enriched for metabolic processes. Genes involved in immune processes and pathways showed a weak upregulation in the transcriptomic analysis. While many genes were newly identified, several other have been previously reported as differentially methylated or expressed in different blood cells from patients with SSc, supporting for their potential dysregulation in SSc.

**Conclusions:** While contrasting with results found in other blood cell types in largely European-descent groups, the results of this study support that variation in DNA methylation and gene expression exists among different cell types and individuals of different genetic, clinical, social, and environmental backgrounds. This finding supports the importance of including diverse, well-characterized patients to understand the different roles of DNA methylation and gene expression variability in the dysregulation of classical monocytes in diverse populations, which might help explaining the health disparities.

## Background

Systemic sclerosis (SSc or scleroderma) is a rare, multisystem, connective tissue disease characterized by cutaneous and visceral fibrosis, immune dysregulation, and vasculopathy. SSc is very heterogeneous, with patients being commonly classified into three subsets based on the pattern of skin involvement: *sine* scleroderma, limited cutaneous SSc (lcSSc), or diffuse cutaneous SSc (dcSSc), the latter having the worse prognosis [1]. SSc is also marked by pronounced gender and ethnic disparities. Similar to other autoimmune diseases, women are four to nine times more likely to have SSc than men [2]. Relative to individuals of European descent, African Americans are more likely to develop SSc [3], have earlier onset of disease, increased disease severity, increased morbidity, earlier mortality, and reduced survival [4-13]. The etiology of SSc and the factors underlying these disparities remain elusive, and African American individuals continue to be underrepresented in research [14].

While having a family history of SSc is a risk factor for developing the disease [15], the low concordance rate of disease between monozygotic twins suggests that epigenetic and/or environmental factors may play a substantial role in SSc pathogenesis [16-18]. Indeed, genetic and epigenetic studies conducted mostly in individuals of European descent have uncovered multiple loci associated with SSc [19, 20]. Variation in DNA methylation across ancestral populations is contributed to by genetic ancestry and environmental factors [21]. Despite the increased disease burden in African Americans and variation in DNA methylation across populations, only two genome-wide differential DNA methylation analyses have been conducted in peripheral blood and skin fibroblasts from SSc patients of African descent [22, 23].

The dysregulation of monocytes in patients with SSc is well established as evidenced by their increased numbers in both peripheral blood and in skin of SSc patients [24-27] and are associated with reduced survival in SSc [27]. African Americans exhibit stronger inflammatory signatures [28-34], including heightened monocyte activation [30, 31]. As recently reviewed, monocytes are associated with altered epigenetic marks in SSc [20]. Notably, histone demethylation and chromatin dysregulation underlie monocyte dysregulation in patients with SSc [16, 35], and contribute to the trans-differentiation of fibroblasts, a key step in the pathogenesis of SSc [36, 37]. Recently, the first transcriptomic analysis of monocytes in patients with SSc revealed great variability of expression patterns across SSc patients that correlated with disease activity outcome measures [38]. The role of DNA methylation and its relationship with gene expression patterns in SSc monocytes has not been previously investigated and studies in health disparity populations are lacking.

Given the dysregulation of monocytes in SSc and the increased prevalence and severity of disease in African Americans, it is important to identify the mechanisms underlying this dysregulation and their potential contribution to the ethnic disparity. In this study, we undertook a systems-level approach, integrating DNA methylation and transcriptional data, to assess the relationship between DNA methylation and gene expression in classical monocytes from African American patients with SSc.

## Results

### Subject characteristics

Given the sex and ethnic disparities in SSc this study focused on African American women, a health disparity population for SSc. All participants were self-reported African American female, and all patients met the 2013 ACR/EULAR classification criteria for SSc [39], most presenting with diffuse cutaneous SSc (dcSSc), interstitial lung disease (ILD), and being on current immunosuppressive therapies. No participants reported current infections or malignancy at the time of study visit. Additional clinical and demographic characteristics of the SSc patients and healthy controls are summarized in Table 1. Classical monocytes (CD14++CD16-) were isolated from the study participants using fluorescence activated cell sorting (FACS).

**Table 1.**
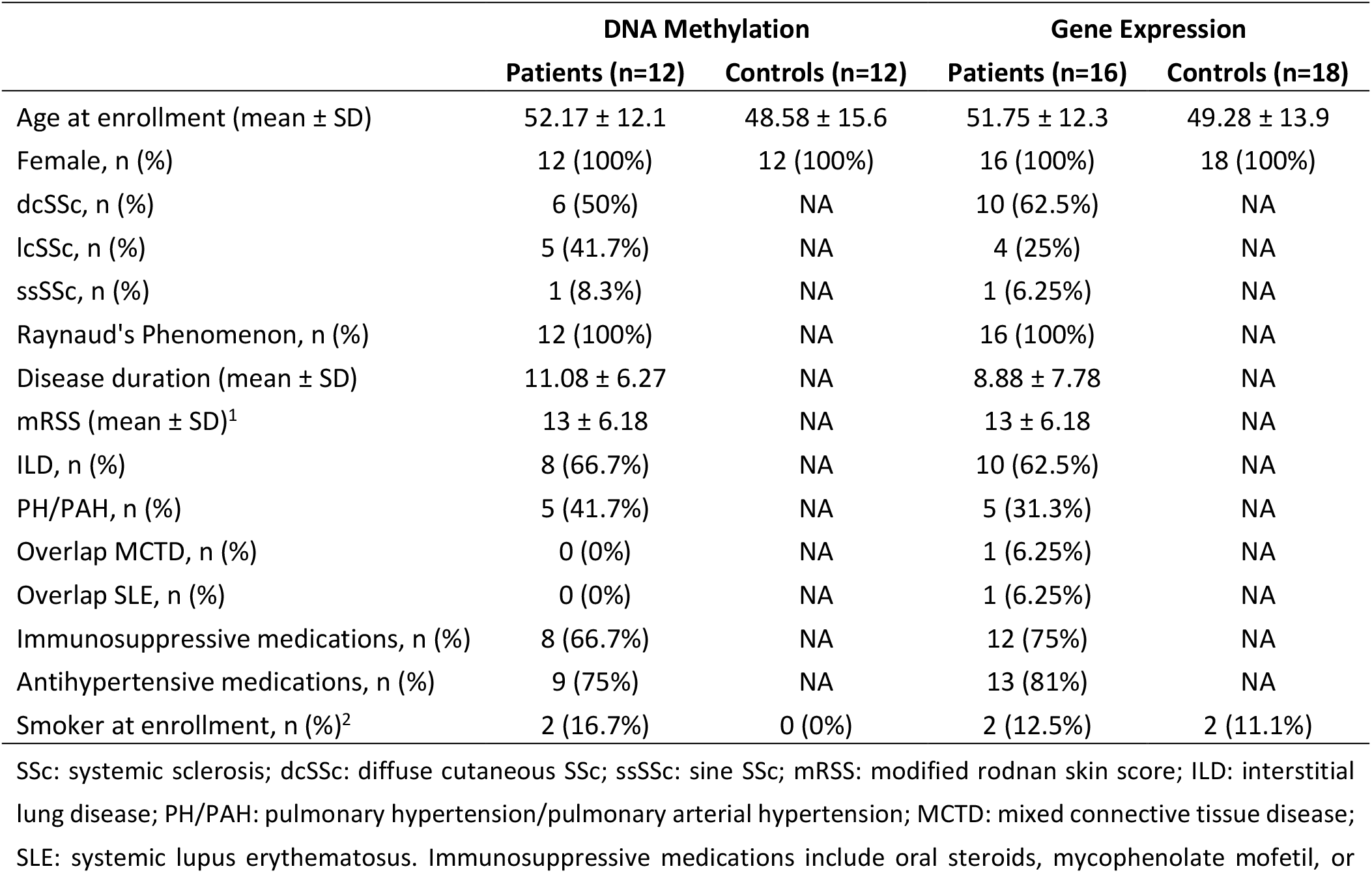
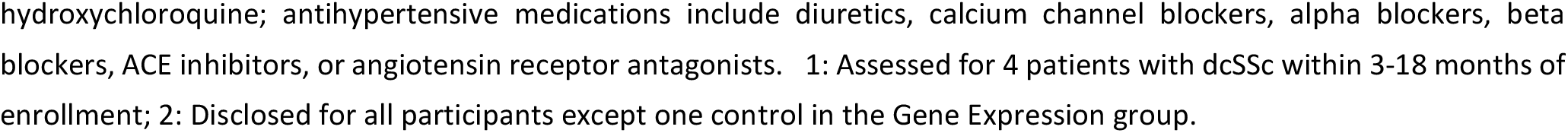
Demographic and clinical characteristics of the study participants.

|  | DNA Methylation |  | Gene Expression |  |
| --- | --- | --- | --- | --- |
|  | Patients (n=12) | Controls (n=12) | Patients (n=16) | Controls (n=18) |
| Age at enrollment (mean ± SD) | 52.17 ± 12.1 | 48.58 ± 15.6 | 51.75 ± 12.3 | 49.28 ± 13.9 |
| Female, n (%) | 12 (100%) | 12 (100%) | 16 (100%) | 18 (100%) |
| dcSSc, n (%) | 6 (50%) | NA | 10 (62.5%) | NA |
| lcSSc, n (%) | 5 (41.7%) | NA | 4 (25%) | NA |
| ssSSc, n (%) | 1 (8.3%) | NA | 1 (6.25%) | NA |
| Raynaud's Phenomenon, n (%) | 12 (100%) | NA | 16 (100%) | NA |
| Disease duration (mean ± SD) | 11.08 ± 6.27 | NA | 8.88 ± 7.78 | NA |
| mRSS (mean ± SD)<sup>1</sup> | 13 ± 6.18 | NA | 13 ± 6.18 | NA |
| ILD, n (%) | 8 (66.7%) | NA | 10 (62.5%) | NA |
| PH/PAH, n (%) | 5 (41.7%) | NA | 5 (31.3%) | NA |
| Overlap MCTD, n (%) | 0 (0%) | NA | 1 (6.25%) | NA |
| Overlap SLE, n (%) | 0 (0%) | NA | 1 (6.25%) | NA |
| Immunosuppressive medications, n (%) | 8 (66.7%) | NA | 12 (75%) | NA |
| Antihypertensive medications, n (%) | 9 (75%) | NA | 13 (81%) | NA |
| Smoker at enrollment, n (%)<sup>2</sup> | 2 (16.7%) | 0 (0%) | 2 (12.5%) | 2 (11.1%) |

SSc: systemic sclerosis; dcSSc: diffuse cutaneous SSc; ssSSc: sine SSc; mRSS: modified rodnan skin score; ILD: interstitial lung disease; PH/PAH: pulmonary hypertension/pulmonary arterial hypertension; MCTD: mixed connective tissue disease; SLE: systemic lupus erythematosus. Immunosuppressive medications include oral steroids, mycophenolate mofetil, or
hydroxychloroquine; antihypertensive medications include diuretics, calcium channel blockers, alpha blockers, beta blockers, ACE inhibitors, or angiotensin receptor antagonists. 1: Assessed for 4 patients with dcSSc within 3-18 months of enrollment; 2: Disclosed for all participants except one control in the Gene Expression group.

### Differentially methylated sites and genes are enriched for metabolic processes

To gain insights into functional and molecular alterations of monocytes in African American patients with SSc, over 850,000 CpG sites were tested for differential methylation between self-reported African American female patients with SSc and controls.

The differences in methylation levels between patients and controls were modest. A total of 19 differentially methylated CpGs (DMCs), which corresponds to 0.002% of all cytosines tested, meet an FDR-adjusted *p*-value < 0.4 (Table 2). The rationale for the FDR setting was guided by the desire to perform a system-level analysis and include as many CpGs sites as possible, as well as previous studies demonstrating that this threshold permits a sensitive analysis at a system level of genes that are relevant to the underlying biology of the trait [40, 41]. A P-P plot of CpG association testing results supports that using –log(*p*) > 4 is a reasonable empirical threshold of significance (Additional file 1: Fig. S1) for systems-level analyses.

**Table 2.** Top differentially methylated CpGs between female African American patients with SSc and controls ranked by absolute effect size.

| CpG | Gene | Chr | Position (kb) | Relation to Island | Control $\beta$ | SSc $\beta$ | Difference | $p$-value | Adjusted $p$-value |
| --- | --- | --- | --- | --- | --- | --- | --- | --- | --- |
| cg22805491 | NIN | 14 | 51,172 | OpenSea | 0.44 | 0.53 | 0.09 | 2.99E-08 | 0.02 |
| cg24073653 | SLC41A2 | 12 | 105,221 | OpenSea | 0.61 | 0.69 | 0.08 | 2.92E-06 | 0.2 |
| cg19933320 | ZNF107 | 7 | 64,125 | N_Shore | 0.33 | 0.4 | 0.08 | 8.20E-06 | 0.35 |
| cg06548512 | LOC728989 | 1 | 146,522 | Island | 0.63 | 0.7 | 0.07 | 1.32E-07 | 0.05 |
| cg00832928 | SELENOT | 3 | 150,329 | Island | 0.61 | 0.68 | 0.07 | 4.58E-07 | 0.11 |
| cg08653580 | CD5 | 11 | 60,862 | OpenSea | 0.66 | 0.73 | 0.07 | 1.21E-06 | 0.16 |
| cg12601237 | ST8SIA6 | 10 | 17,429 | OpenSea | 0.71 | 0.78 | 0.07 | 3.69E-06 | 0.21 |
| cg14115740 | FANCC | 9 | 98,055 | Island | 0.57 | 0.64 | 0.07 | 4.74E-06 | 0.24 |
| cg22167498 | RAB11B-AS1 | 19 | 8,451 | N_Shelf | 0.57 | 0.62 | 0.06 | 1.00E-06 | 0.16 |
| cg23596249 | MARCHF1 | 4 | 165,110 | Island | 0.51 | 0.57 | 0.06 | 1.85E-06 | 0.19 |
| cg13704629 | DCAF4 | 14 | 73,396 | S_Shelf | 0.56 | 0.62 | 0.06 | 2.73E-06 | 0.2 |
| cg11652329 | P2RX6P | 22 | 21,399 | N_Shore | 0.61 | 0.67 | 0.05 | 4.74E-06 | 0.24 |
| cg26715639 | S100A11 | 1 | 151,986 | OpenSea | 0.6 | 0.65 | 0.05 | 6.43E-06 | 0.31 |
| cg23493751 | CCR3 | 3 | 46,205 | OpenSea | 0.75 | 0.8 | 0.05 | 7.64E-06 | 0.34 |
| cg08288426 | ZSCAN29 | 15 | 43,651 | OpenSea | 0.8 | 0.76 | -0.04 | 1.51E-06 | 0.17 |
| cg18507060 | OAS3 | 12 | 113,399 | OpenSea | 0.77 | 0.81 | 0.04 | 5.48E-07 | 0.11 |
| cg10044900 | CFAP44 | 3 | 113,058 | OpenSea | 0.76 | 0.8 | 0.04 | 2.55E-06 | 0.2 |
| cg14929421 | ACTR3BP2 | 2 | 92,318 | OpenSea | 0.52 | 0.55 | 0.03 | 2.80E-06 | 0.2 |
| cg04331667 | CCDC71L | 7 | 106,171 | OpenSea | 0.8 | 0.83 | 0.03 | 3.21E-06 | 0.2 |

CpGs are shown, along with nearest gene, annotation, averaged methylation levels ( $\beta$ ), methylation difference, unadjusted and adjusted $p$ -values.

In addition to CpGs near several pseudogenes, top differentially methylated CpGs included those near the genes that encode the centrosomal protein ninein (NIN, aka Glycogen Synthase Kinase 3 Beta-Interacting Protein), the selenoprotein T (SELENOT), the synthetase OAS3, or the melanoprotein T-Cell Surface Glycoprotein CD5 (Table 2; Additional file 1: Figs. S2 and S3).

We sought to investigate any potential enrichment (or conversely, underrepresentation) of DMCs in defined genomic regions (Fig. 1). Among the top 100 DMCs, there was an overrepresentation of DMCs in exon boundaries (OR = 4.9, *p* < 0.0001), while there was a depletion of DMCs in the vicinity of transcription start sites (OR = 0.2, *p* < 0.005).

**Fig. 1.**
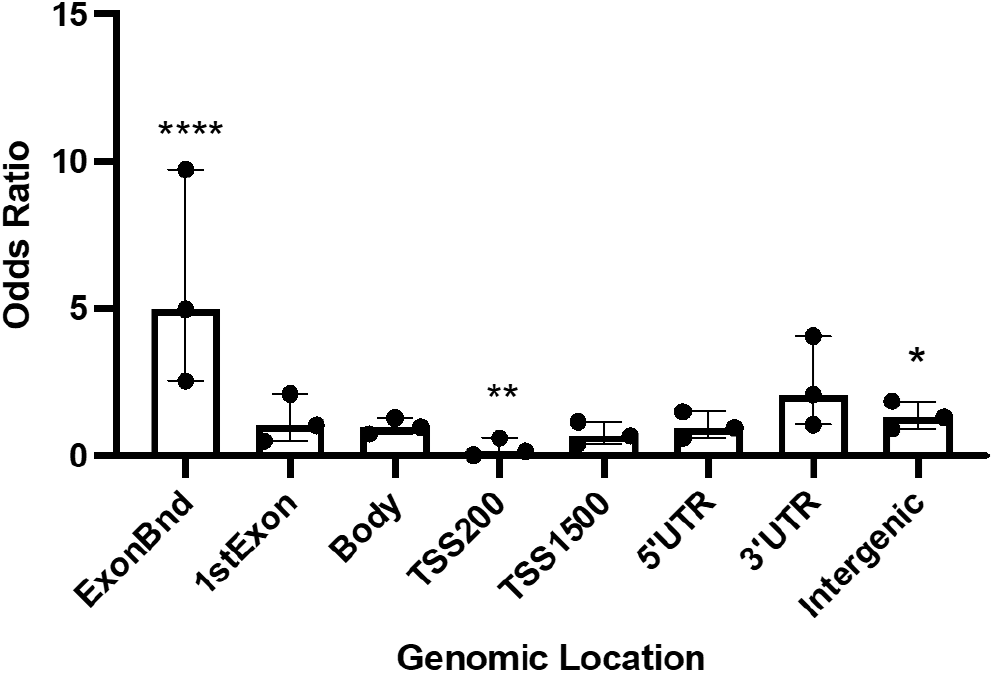
Genomic location of the top 100 differentially methylated CpGs (DMC). Odds ratio (OR), 95% confidence intervals (CI), and *p*-values were computed against the general distribution of the CpGs using GraphPad Prism v9. Error bars represent the 95% CI. OR indicates the enrichment or depletion of DMCs in each region. * *p*<0.05; ** *p*<0.005; **** *p*<0.0001. TSS: transcription start site. TSS200: 0–200 bases upstream of the transcriptional start site (TSS). TSS1500: 200–1500 bases upstream of the TSS.

To better understand the chromatin context and functional role underlying the disease-associated CpG sites, we performed integrative epigenomics analyses using the eFORGE 2.0 framework [42-44] to assess whether these SSc-associated CpGs reside within regulatory regions across the genome in diverse tissues and cell types. Using the top 100 DMCs associated with SSc showed an enrichment of H3K9me3 (a histone mark associated with heterochromatin regions, important for repressing repetitive elements, non-coding portions of the genome, and silencing lineage-inappropriate genes) in several fetal cells (blue in Fig. 2), and an enrichment of H3K36me3 (a transcription-associated histone mark important in maintaining gene expression stability and regulation of DNA damage repair) in several blood, stem and fetal cells (pink in Fig. 2). Overall, these findings suggest that most epigenetic changes are present in non-transcribed regions.

**Fig. 2.**
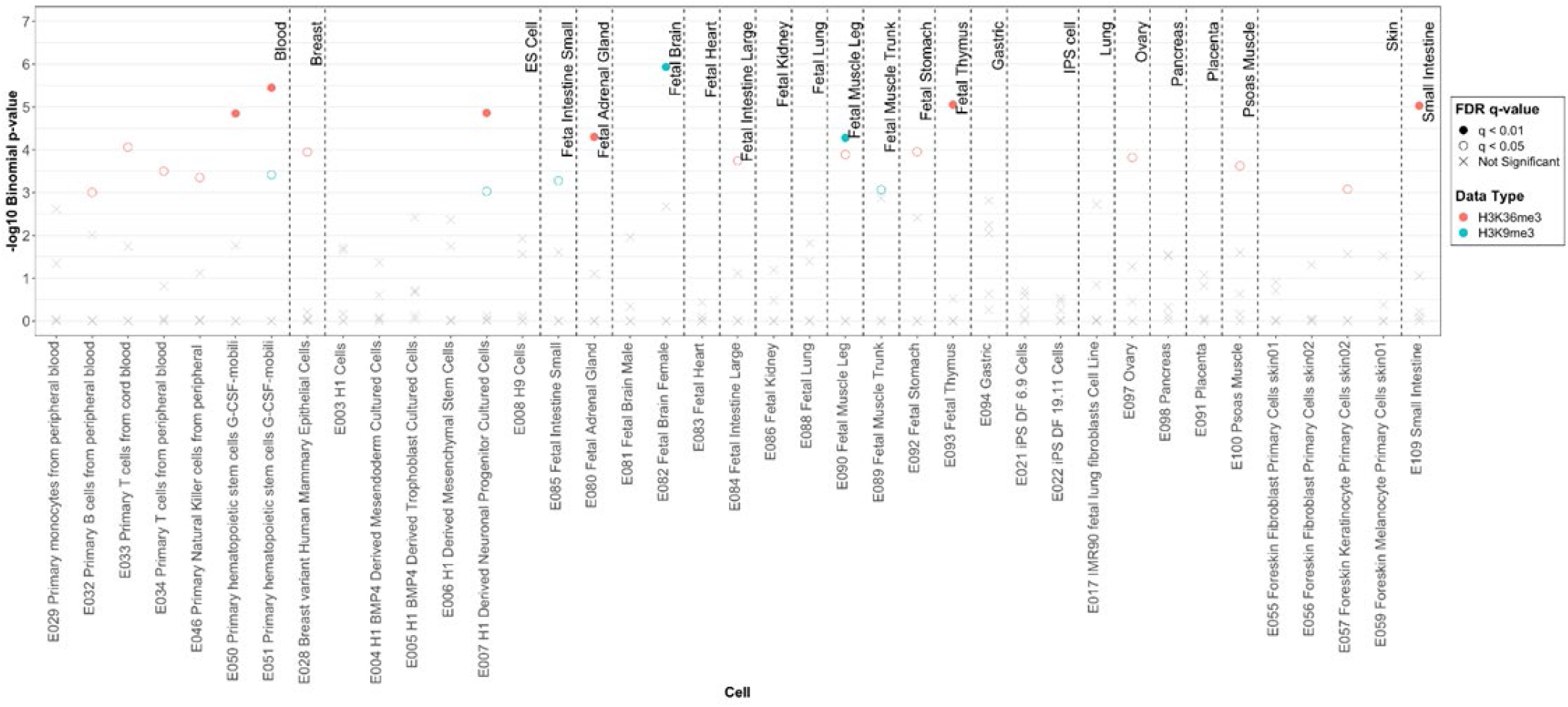
Enrichment of differentially methylated CpGs in H3K9me3 and H3K36me3 histone marks among various cell and tissue types using Roadmap Epigenomics project data. Statistically significant enrichment outside the 99.9th percentile (−log10 binomial p-value ≥ 3.38) is colored red on the vertical axis.

We first used the DAVID Functional Annotation Tool 6.8 [45, 46] to uncover the biological significance of the genes in the regions of the differentially methylated cytosines shown in Table 2. Although not significant, the top Gene Ontology (GO) terms were related to Metabolic Processes (GO:0008152; *p* = 0.4), driven by OAS3, DCAF4, FANCC, S100A11, ST8SIA6, RNF24, SELENOT, ZSCAN29, and ZNF107. Close to half of the genes harboring DMC are phosphoproteins: OAS3, CCR3, S100A11, CFAP44, CCDC71L, SLC41A2, ZSCAN29, CD5, NIN.

Next, we used *Enrichr* [47, 48] to explore the Biological Processes and KEGG pathways associated with the genes harboring the top DMCs in Table 2. As shown in Fig. 3, the top Biological Processes were related to metabolic processes, including those of RNA (GO:0060700, *p* = 4.9E-03 (driven by the ribonuclease OAS3)) and protein (GO:0032069, *p* = 5.9E-03 (driven by the selenocysteine SELENOT)) metabolic processes, as well as hormone secretion (GO:0060124, *p* = 5.9E-03 (driven by OAS3 and SELENOT)). No pathways showed significant enrichment.

**Fig. 3.**
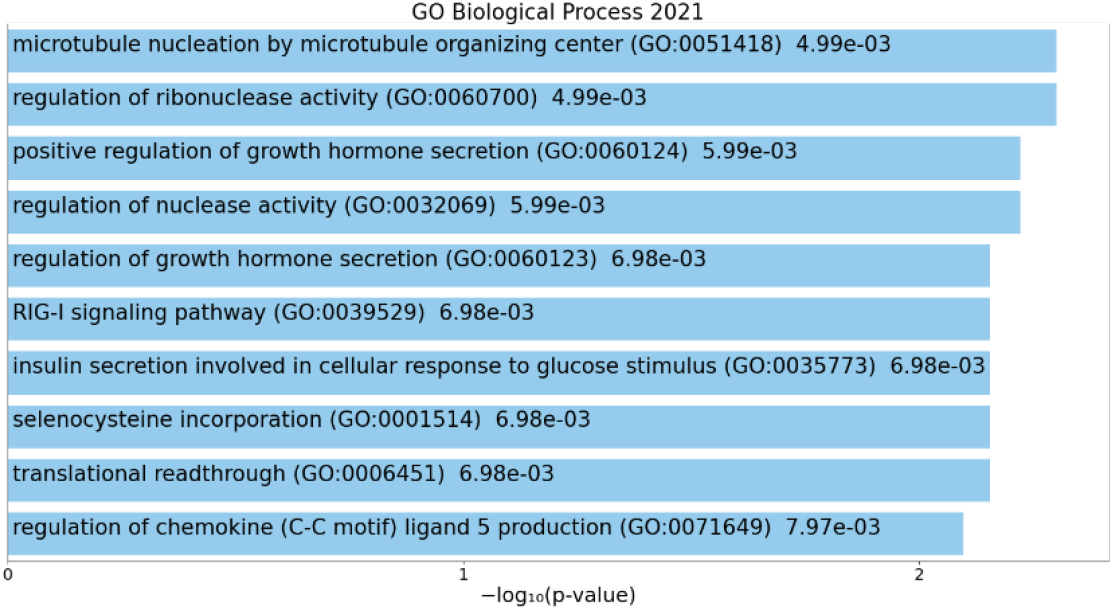
Metabolic process enrichment among the genes harboring the top DMCs. The x-axis shows the − log10(*p*-value) of the Biological Process enrichment obtained from Enrichr. The length of the bar represents the significance of that specific term.

### Differentially expressed genes are enriched for metabolic processes

Differential gene expression analysis revealed a total of 1,272 transcripts differentially expressed between African American female patients with SSc and controls at an FDR-corrected *p* < 0.4 (Additional file 1: Fig. S4). These differentially expressed transcripts correspond to 5.0% of all 25,369 transcripts tested. The rationale for the FDR setting was the same as described above for the DNA methylation analysis. The top differentially expressed transcripts are shown in Table 3.

**Table 3.**
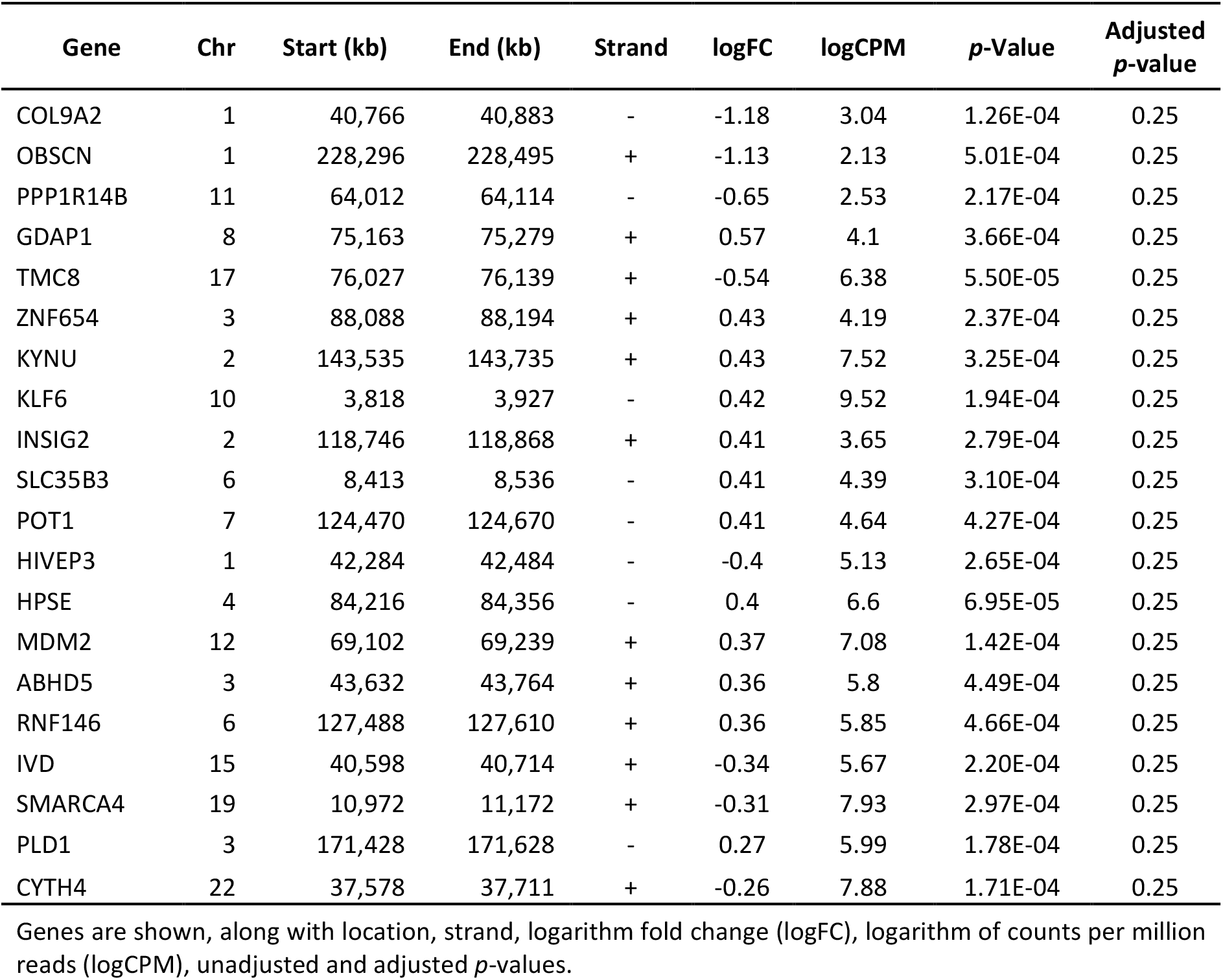
Top 20 differentially expressed transcripts between female African American patients with SSc and controls ranked by absolute effect size.

| Gene | Chr | Start (kb) | End (kb) | Strand | logFC | logCPM | *p*-Value | Adjusted *p*-value |
| --- | --- | --- | --- | --- | --- | --- | --- | --- |
| COL9A2 | 1 | 40,766 | 40,883 | - | -1.18 | 3.04 | 1.26E-04 | 0.25 |
| OBSCN | 1 | 228,296 | 228,495 | + | -1.13 | 2.13 | 5.01E-04 | 0.25 |
| PPP1R14B | 11 | 64,012 | 64,114 | - | -0.65 | 2.53 | 2.17E-04 | 0.25 |
| GDAP1 | 8 | 75,163 | 75,279 | + | 0.57 | 4.1 | 3.66E-04 | 0.25 |
| TMC8 | 17 | 76,027 | 76,139 | + | -0.54 | 6.38 | 5.50E-05 | 0.25 |
| ZNF654 | 3 | 88,088 | 88,194 | + | 0.43 | 4.19 | 2.37E-04 | 0.25 |
| KYNU | 2 | 143,535 | 143,735 | + | 0.43 | 7.52 | 3.25E-04 | 0.25 |
| KLF6 | 10 | 3,818 | 3,927 | - | 0.42 | 9.52 | 1.94E-04 | 0.25 |
| INSIG2 | 2 | 118,746 | 118,868 | + | 0.41 | 3.65 | 2.79E-04 | 0.25 |
| SLC35B3 | 6 | 8,413 | 8,536 | - | 0.41 | 4.39 | 3.10E-04 | 0.25 |
| POT1 | 7 | 124,470 | 124,670 | - | 0.41 | 4.64 | 4.27E-04 | 0.25 |
| HIVEP3 | 1 | 42,284 | 42,484 | - | -0.4 | 5.13 | 2.65E-04 | 0.25 |
| HPSE | 4 | 84,216 | 84,356 | - | 0.4 | 6.6 | 6.95E-05 | 0.25 |
| MDM2 | 12 | 69,102 | 69,239 | + | 0.37 | 7.08 | 1.42E-04 | 0.25 |
| ABHD5 | 3 | 43,632 | 43,764 | + | 0.36 | 5.8 | 4.49E-04 | 0.25 |
| RNF146 | 6 | 127,488 | 127,610 | + | 0.36 | 5.85 | 4.66E-04 | 0.25 |
| IVD | 15 | 40,598 | 40,714 | + | -0.34 | 5.67 | 2.20E-04 | 0.25 |
| SMARCA4 | 19 | 10,972 | 11,172 | + | -0.31 | 7.93 | 2.97E-04 | 0.25 |
| PLD1 | 3 | 171,428 | 171,628 | - | 0.27 | 5.99 | 1.78E-04 | 0.25 |
| CYTH4 | 22 | 37,578 | 37,711 | + | -0.26 | 7.88 | 1.71E-04 | 0.25 |

Genes are shown, along with location, strand, logarithm fold change (logFC), logarithm of counts per million reads (logCPM), unadjusted and adjusted *p*-values.

The top differentially expressed genes (DEGs) include the collagen *COL9A2*, the endoplasmic reticulum transmembrane channel-like *TMC8* gene, the heparinase *HPSE*, the proto-oncogene nuclear ubiquitin ligase *MDM2*, the vesicular trafficking cytohesin *CYTH4*, the phospholipase *PLD1*, and the Kruppel-like transcription factor *KLF6* (Table 3).

Gene Ontology analysis of the top transcripts (Table 3) using DAVID 6.8 showed many genes that participate in metabolic processes (GO:004423; *p* = 2.70E-02 (KLF6, MDM2, SMARCA4, ABHD5, GDAP1, HPSE, HIVEP3, INSIG2, IVD, KYNU, OBSCN, PLD1, POT1, PPP1R14B, RNF146, SLC35B3, ZNF654)), especially catabolic processes (GO:0044248; *p* = 2.0E-02 (MDM2, ABHD5, HPSE, IVD, KYNU, PLD1, RNF146), and sulfur compound metabolic process (GO:0006790; *p* = 8.0E-03 (GDAP1, HPSE, KYNU, SLC34B3)).

*Enrichr* analysis of the top differentially expressed genes in Table 3 revealed an enrichment of several biological processes related to metabolic processes (e.g., GO:0006654, *p* = 1.0E-03), as well as an enrichment of the KEGG endocytosis pathway (*p* = 1.9E-03) (Fig. 4A and B). Relaxing the threshold and including the 450 transcripts with FDR-corrected *p* < 0.3 in the comparison between cases and controls revealed an involvement of immune biological processes and pathways (Fig. 4C and D).

**Fig. 4.**
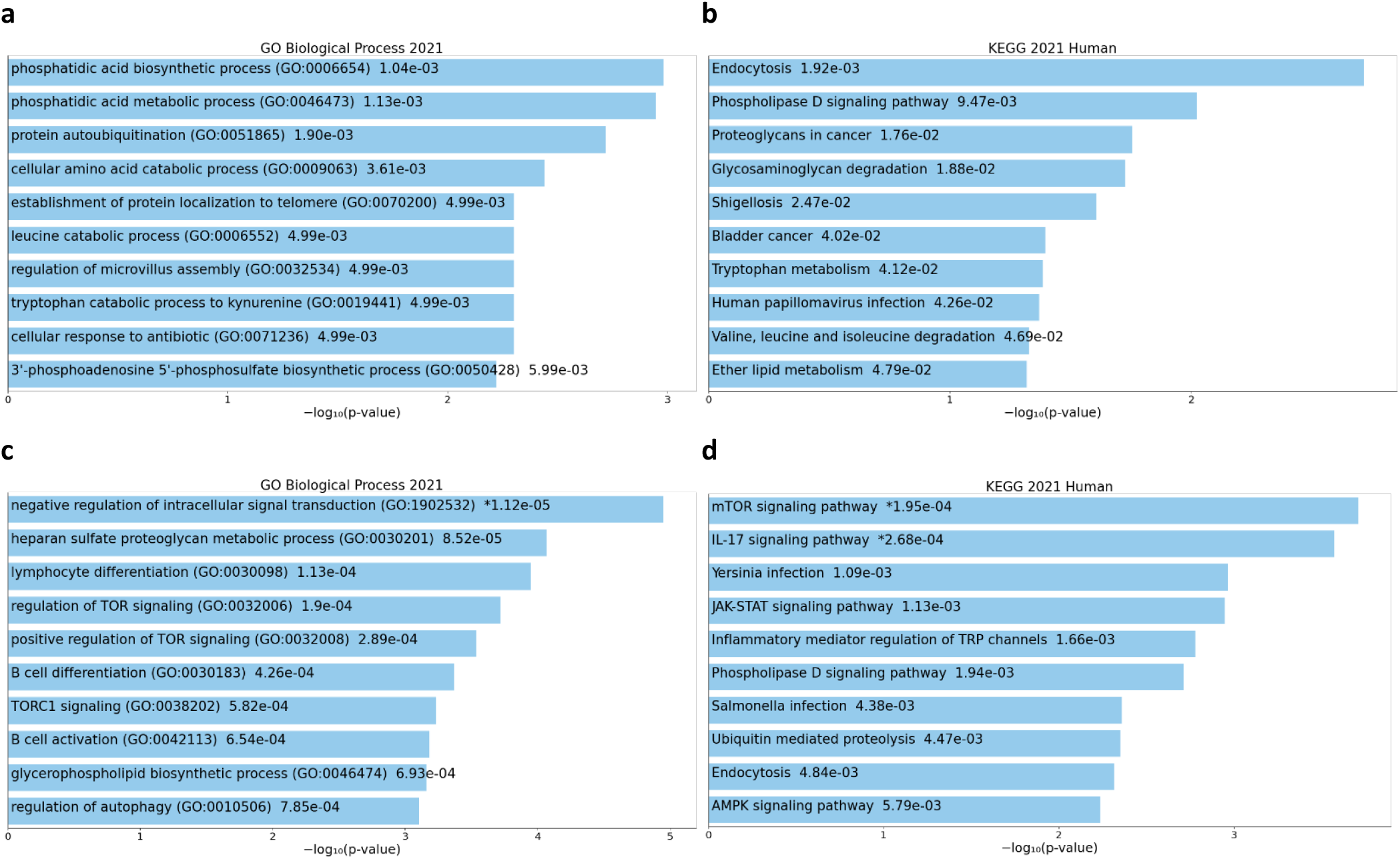
Metabolic and immune process and pathway enrichment among differentially expressed genes. Biological processes (**a**) and KEGG pathways (**b**) enriched for the top 20 DEG; biological processes (**c**) and KEGG pathways (**d**) enriched for the top 450 DEG. The x-axis shows the − log10(*p*-value) of enrichment obtained from Enrichr. The length of the bar represents the significance of that specific term.

### Expression quantitative trait methylation (eQTM) analysis

To investigate the potential mechanistic relationship between DNA methylation and gene expression variation in classical monocytes, we leveraged RNA-sequencing data from the same individuals and computed an expression quantitative trait methylation (eQTM) analysis to identify CpGs associated with changes in gene expression. Table 4 shows the top eQTM loci, which include the transcriptional repressor homolog *PCGF1*, the RNA helicase *DDX27*, and the splicing factor *SF3B2*. About half the eQTM loci showed a negative correlation between DNA methylation and gene expression levels, while the other half displayed a positive correlation (Table 4).

**Table 4.**
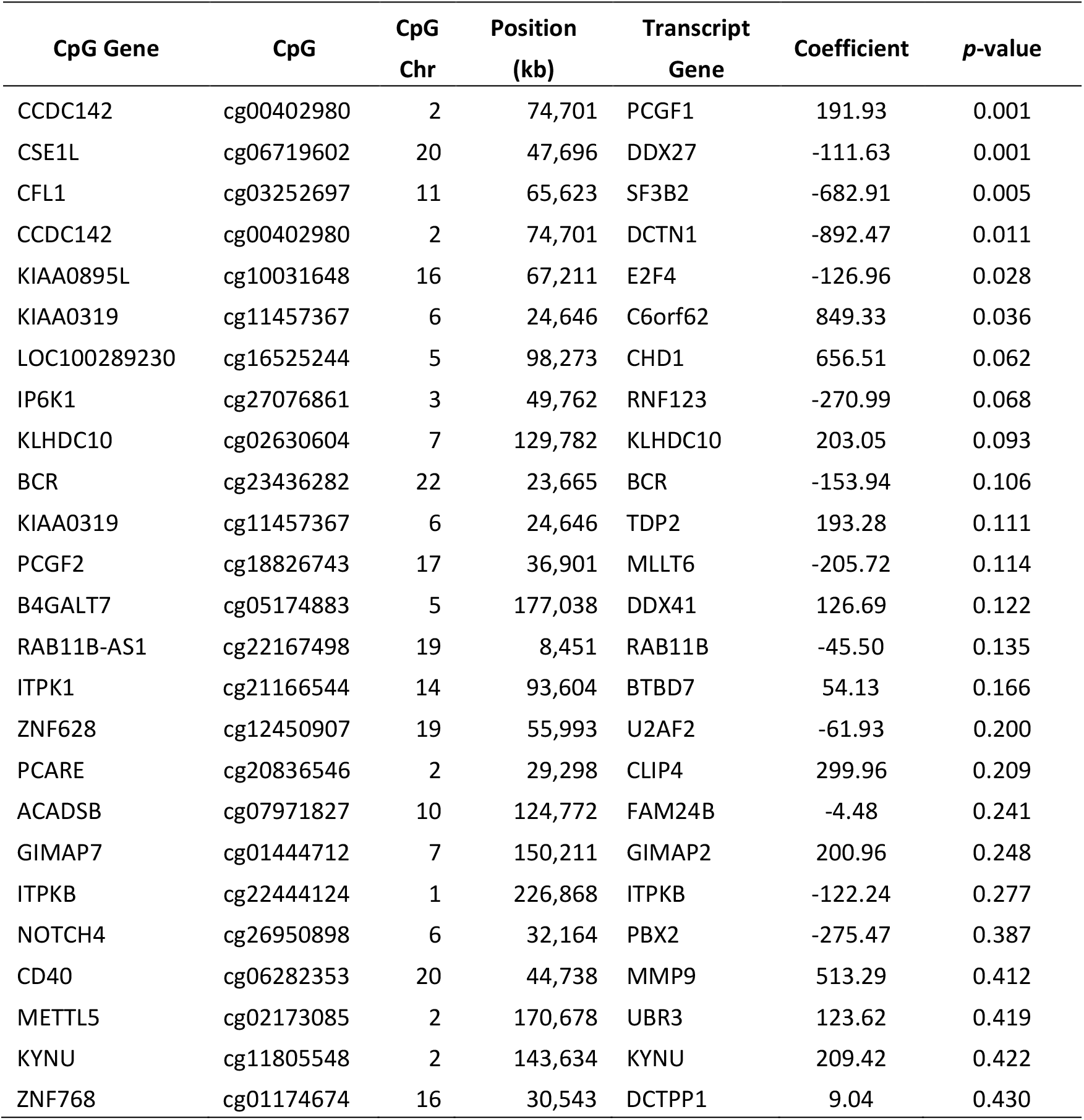
Top 25 eQTM loci

These 25 genes whose expression is associated with DMCs are enriched for cellular metabolic processes according to DAVID (GO:0044237, *p* = 6.50E-03 (driven by BCR, DDX27, DDX41, E2F4, MLLT6, PBX2, RAB11B, U2AF2, CHD1, DCTPP1, ITPKB, KYNU, MMP9, PCGF1, RNF123, SF3B2, TDP2, UBR3)). In addition to microtubule cytoskeleton organization processes, other metabolic processes like RNA splicing and processing were also unveiled by Enrichr (Fig. 5). The top KEGG Pathway was vasopressin-regulated water reabsorption (Fig. 5).

**Fig. 5.**
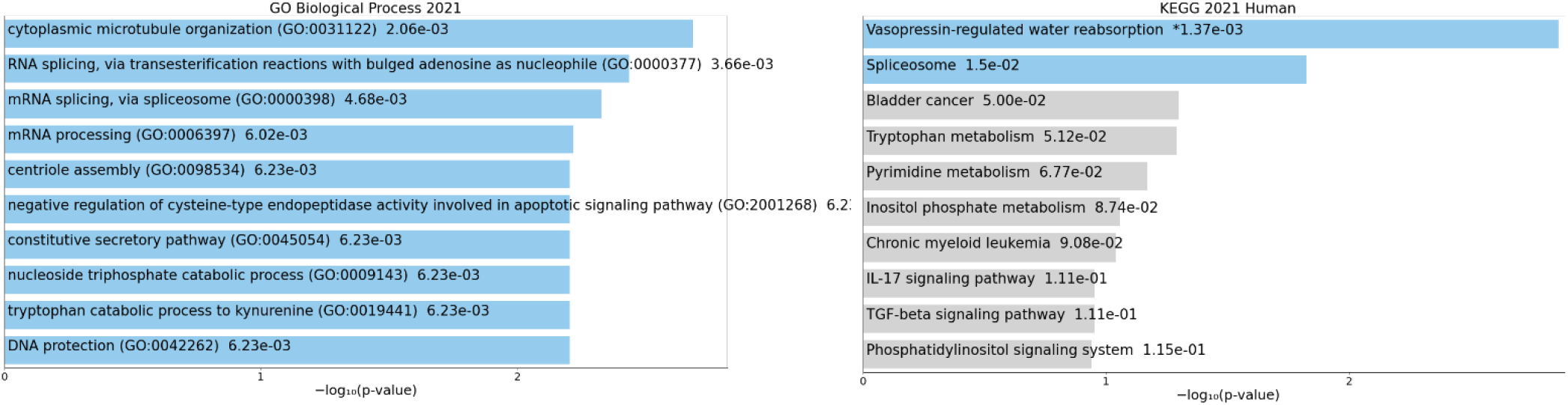
Biological processes (**a**) and KEGG pathways (**b**) enrichment among genes associated with eQTM loci. The x-axis shows the − log10(*p*-value) of enrichment obtained from Enrichr. The length of the bar represents the significance of that specific term. Terms displayed in gray are not significant.

## Discussion

This study is to our knowledge the first to evaluate and integrate analysis of DNA methylation and gene expression in classical monocytes from African American patients with SSc on a genome-wide scale. We show modest differences in DNA methylation and gene expression between patients and controls, and an enrichment of genes involved in metabolic processes.

The differential methylation analysis showed that the top DMCs were enriched for H3K9me3 and H3K36me3, markers associated with heterochromatic regions in several fetal tissues. This is consistent with the recent finding that several tissue-specific repressed genomic regions are enriched for disease-associated GWAS variants, and suggests that DMC may also have tissue-specific effects in repressive regions [44]. Previous analyses of DNA methylation in other blood cell types and in largely European-descent groups [22, 49-54] report a larger difference in DNA methylation patterns between cases and controls, and an enrichment of genes involved in immune and inflammatory processes. In our study, only *OAS3* and *CD5* have been previously reported as differentially methylated in CD4+ T cells from Spanish patients with SSc [54]. OAS3 is an interferon-induced, dsRNA-activated antiviral enzyme which plays a role in cellular innate antiviral response and the immune response to the interferon pathway [55, 56]. Its expression can be increased in SSc patients [57]. CD5 is a type-I transmembrane glycoprotein found on the surface of thymocytes, T lymphocytes, and a subset of B lymphocytes.

CD5 is upregulated in B cells from patients with SSc [58]. Among other genes harboring top DMCs, the centrosomal protein NIN has a role in promoting angiogenesis [59], which is dysregulated in SSc. Antibodies against NIN are present in sera from several patients with SSc and other autoimmune disorders [60], which further supports a role for NIN in SSc. Interestingly, NIN may also be regulated by GSK-3β, a key regulator of the canonical Wnt signaling in fibroblasts whose inhibition results in fibroblast activation and increased release of collagen [61].

Multiple analyses of gene expression patterns in different blood cell types from mostly European-derived populations consistently report a prominent upregulation of genes involved in immune and inflammatory processes in SSc patients [38, 62-75]. This includes the only analysis of classical monocytes to date [38], and contrasts with the weaker upregulation of inflammatory and immune genes we observed in classical monocytes from African American SSc patients. Multiple DEGs found in our analysis have been previously reported as differentially expressed in different blood cells from patients with SSc, including the endoplasmic reticulum transmembrane channel-like *TMC8* [76], the collagen *COL9A2* [67], the Kruppel-like transcription factor *KLF6* [38, 62, 76], the zinc finger ZNF654 [76], the insulin-Induced INSIG2 [76], the transcription regulator SMARCA4 [76], the kynureninase *KYNU* [38, 63, 64, 69], the telomere regulator POT1 [76], and the hydrolase *ABHD5* [64, 69]. However, when we focus on classical monocytes, our results are consistent in significance and directionality of effect to those reported by Makinde and colleagues for *HPSE, CYTH4, KLF6, KYNU, C6orf62, and TUBB4B* [38], providing support for their potential dysregulation in SSc.

The variable correlation between methylation patterns and gene expression is well established, being either positive or negative, and being tissue and context specific, in that the local DNA sequence and genomic features largely account for local patterns of methylation [34, 77-83]. In our eQTM analysis we found that about half of the DMCs have a positive and half a negative correlation with gene expression levels. Several of the genes whose expression was associated with eQTM have been previously reported as differentially expressed in different blood cells from patients with SSc, lending further support for their dysregulation in SSc patients. These include DDX27 [76], C6orf62 [76], CHD1 [66], BCR [76], TDP2 [76], MLLT6 [76], RAB11B [76], CLIP4 [76], PBX2 [76], MMP9 [71], and KYNU [38, 63, 64, 69].

Our results provide support for the involvement of dysregulated metabolic processes in SSc, consistend with previous studies reporting that dysregulated metabolism is associated with SSc [84]. Different metabolic perturbations are expressed in different patients, reflecting the clinical heterogeneity of SSc [84]. Our enrichment of genes involved in metabolic processes contrasts with the prominent enrichment of genes involved in immune and inflammatory processes consistently reported in both DNA methylation [22, 49-53, 76] and gene expression analyses in multiple blood cell populations and in largely European-descent groups [38, 62-75]. This is not surprising, given that previous studies have focused on blood (a heterogeneous tissue) or lymphoid cell populations, while our focus was on a myeloid cell population. The distinct racial category of the participants might appear as another explanation for this difference. However, we caution against a simplistic explanation solely based on self-reported race. Race is an imperfect proxy for social determinants of health such as racism and discrimination, economic stability, healthcare access and quality, education access, and environmental exposures. These social and environmental determinants are differentially experienced across groups and geography, resulting in health disparities. We postulate that these sociocultural factors experienced by our study participants are one of the reasons underlying our results. The other reason that can explain the lack of replication across populations is genetic ancestry. Differences in DNA methylation are known to exist between individuals of African and European ancestry [21, 28, 34, 85-89], due to both variation in genetic ancestry and environmental factors [21]. These differences help explain the new findings and minimal overlap with previous reports.

Several other reasons and important limitations of this study might underlie the modest differences in DNA methylation and gene expression we observed in our study focused on classical monocytes from self reported Black women. First, although this study is the first to evaluate DNA methylation and gene expression patterns in monocytes of African Americans, the sample size is very small. This study has virtually no power to detect the DNA methylation differences herein reported, but is powered to detect a reasonable fraction of the differentially expressed transcripts between cases and controls. Nevertheless, with 16 total SSc cases, it is comparable to sample sizes from previous studies that assessed genome-wide DNA methylation and gene expression primarily in European Americans [16, 38, 90, 91]. Also, our results are consistent with the heightened monocyte activation previously reported in African American patients with SSc [30, 31].

Second, we do not have DNA available on these study participants to allow us to estimate their genetic ancestries, and this study relies on self-reported race. Although we observed no evidence of population structure and adjusted for population stratification, to mitigate against the limitations of this study it is essential that future studies genotype and including multiple African groups to fully capture African genetic diversity.

Third, SSc is a rare disease with a prevalence of only 49,000 US adults [92], there is currently no existing cohort or repository of samples from African American patients that can be leveraged to enable large-scale studies, to replicate, and to validate our results. Given our small sample size, we tried to balance our desire to increase discovery at the cost of additional loci being false positive results, which is a reasonable premise to pursue systems-level analyses.

Fourth, most of the patients in our study presented with more severe SSc and were on current immunosuppressive therapies that can impact their epigenetic and transcriptional patterns. Although the modest sample size of this study precludes a robust statistical adjustment for immunosuppressive drug use, and these medications affect the methylation levels of several CpGs, we also show that their use does not cause a substantial bias in our study (Additional file 1: Figs. S6 and S7). Interestingly, in their transcriptional analysis of classical monocytes in SSc patients, Makinde *et al* [38] which included participants of different self-reported racial groups, note the substantial variability in the transcriptional profiles of the patients, with several patients, especially those on current immunosuppressive therapies, more closely resembling controls. This observation is consistent with our results where African American patients, who tend to have more severe disease and be on immunosuppressants, show modest differences in gene expression relative to controls.

Fifth, and inherent to all epigenomic studies, we cannot exclude the possibility of reverse causation, or whether the DNA methylation and gene expression changes are a cause or an effect of SSc. Future longitudinal studies will help to elucidate the role of DNA methylation in disease etiology. Sixth, it is possible that the DNA methylation changes are due to genetic variation, but we lack genotypic data on these samples. Seventh, SSc is a very heterogeneous disorder, which can also explain the inconsistent and sometimes disparate results observed in different studies, including the lack of association between gene expression signatures and clinical characteristics [e.g., [93, 94]]. Similarly, African Americans are a heterogeneous category of individuals with diverse cultural and genetic backgrounds [95].

Finally, we recognize that it is difficult to account for all lifestyle factors that could affect DNA methylation [18]. Unless studies account for all the genetic, clinical, demographic, behavioral, social, and environmental characteristics of their participants, limited reproducibility is not unexpected. Future studies including diverse individuals with measures of genetic ancestry as well as environmental and social determinants of health responsible for the health disparities will ensure the validity and relevance of these findings for patients of all backgrounds. Despite these limitations, the findings in this study further support the need to continue to investigate the regulatory architecture of different cell types in diverse SSc patients.

## Conclusions

Our study suggests that classical monocytes from African American female patients with SSc display modest changes in DNA methylation and gene expression relative to healthy controls. The genes associated with the DMCs, DEGs, as well as eQTM loci, show an enrichment of metabolic processes, and only a weak upregulation of immune processes and pathways. These differences relative to previous reports of differential methylation and gene expression in patients with SSc epitomize the clinical, biological, social, and environmental heterogeneity of SSc patients. This study underscores the importance of research in patients with diverse clinical and sociodemographic characteristics, and of integrating genetic and social factors, to enable a thorough understanding of the different roles of DNA methylation and gene expression variability in the dysregulation of classical monocytes in different populations.

## Methods

### Subjects

A total of 34 self-reported Black or African American females were recruited for this study: 16 patients with SSc and 18 healthy controls. All patients met the 2013 ACR/EULAR classification criteria for SSc [39]. Cases and controls were age-balanced within five years.

### Classical monocyte isolation

Peripheral blood (40 ml) was drawn by venipuncture into EDTA tubes and stored at 4°C overnight. Peripheral blood mononuclear cells (PBMCs) were isolated using Sep-Mate tubes and Lymphoprep (Stem Cell Technologies, Cambridge, MA, USA) according to manufacturer guidelines. After isolation, any residual red blood cells were depleted via red blood cell lysis (144 mM NH_4_Cl and 17 mM Tris, pH 7.6), washed twice with 1X PBS, and stored at −80°C in 90% Fetal Bovine Serum (FBS)/10% dimethyl sulfoxide (DMSO). After thawing, PBMCs were stained with CD-14 Brilliant violet 421 (1:100) and CD-16 Brilliant violet 605 (1:100). Cells were incubated with antibodies for 30 min on ice in the dark. Both antibodies were purchased from Biolegend (San Diego, CA, USA). Viability was assessed using Near-infrared Live/Dead Fixable Dead Cell stain (Life Technologies, Carlsbad, CA, USA) at a concentration of 50 μl/ million cells. CD14++/CD16-cells were collected with a FACSAria III cell sorter (BD Biosciences, San Jose, CA, USA).

### MethylationEPIC Assays, Quality Control and Batch Normalization

DNA was extracted using DNeasy kits (Qiagen, Germantown, MD, USA) according to manufacturer protocols from classical monocytes (CD14++CD16-) isolated from 12 female African American SSc patients and 12 female African American controls. DNA methylation was assessed using Illumina’s MethylationEPIC BeadChip (Illumina, San Diego, CA, USA). 500 ng of each sample was bisulfite converted using an EZ DNA Methylation Kit (Zymo, Tustin, CA, USA), amplified, hybridized and imaged. DNA methylation data for over 850,000 CpGs were generated per sample and preprocessed using GenomeStudio in the form of beta values, which is the estimate of the proportion of methylation in a cell population. GenomeStudio also produced detection p-values, which is the probability that the intensity is due to background noise rather than a true signal. After filtering out background noise, ComBat [96] was used as an empirical Bayes approach to correct for differences between batches. A single array containing 12 samples and approximately 20,000 CpGs separated at random was used to describe a batch to be normalized on our in-house, high-performance computing cluster at the HudsonAlpha Institute for Biotechnology. After correcting for batch effects, beta values between the two probe types, Infinium I & Infinium II, were normalized by using the equation: *Beta*_*Type II Probe*_ *= γ + β*_1_ *\* Beta*_*Type II Probe*_ *+ β*_2_ *\* Beta*_*Type II Probe*_^2^ The intercept and beta coefficients were calculated by fitting a second order polynomial to beta values from paired Infinium I and Infinium II CpGs that were within 50 bp of each other as described previously [97].

Only good quality samples (successful probe ratio >0.1) were included. CpG probes with detection p-values over 0.01 were removed. The R package ChAMP [98] was used to exclude non-CpG probes, probes that have been previously reported to bind to multiple locations [99], and probes with a bead count less than 3 in ≥ 5% of samples per probe.

Given the lack of genetic ancestry estimates we used EPISTRUCTURE, a method for the inference of ancestry from methylation data that relies on reference data in which both genotype and methylation data are available [100]. The PCA plot generated using EPISTRUCTURE shown in Additional file 1: Fig. S5 shows no evidence of population structure. Smoking is known to affect methylation across the genome, therefore CpGs known to be associated with smoking were removed prior to analysis [101]. After pre-processing and filtering of the methylation data, 817,938 CpGs remained for downstream analysis. Genome-wide data methylation analysis was then performed using the R statistical suite (version 3.6.3) [102]

### Genome-wide DNA Methylation Regression Analysis

Linear regression analyses were performed at each CpG using the stats package in R [102] to test whether that particular CpG was associated with SSc by examining DNA methylation differences between patients and controls. Principal components were calculated using the built-in R function prcomp() from the stats package and used in our regression model to correct for unknown potential sources of variance such as admixture. In our model, disease status was considered the random effect, and the first two principal components were considered fixed; due to age being known to influence DNA methylation, it was also placed as a covariate:

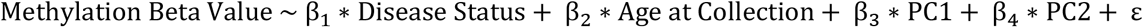

The associated CpG p-values were then corrected using the Benjamini-Hochberg False Discovery Rate (FDR) method. Given the paucity of differentially methylated cytosines identified with FDR-corrected *p* < 0.05, cytosines with an FDR-corrected *p* < 0.4 are reported. The rationale for the FDR setting was guided by the desire to perform a system-level analysis and include as many CpGs sites as possible, as well as previous studies demonstrating that this threshold permits a sensitive analysis at a system level of genes that are relevant to the underlying biology of the trait [40, 41]. The premise that most CpGs have sufficient quality, coupled with the desire to increase discovery at the cost of additional loci being false positive results, is a reasonable premise for intitial studies to pursue pathway and network analyses. To further select an empirical threshold for significance using a data-informed approach, we generated the P-P plot to see the point of departure where the CpGs start deviating from the null (Additional file 1: Fig. S1). As shown in Additional file 1: Fig. S1, fitting a linear regression line for the theoretical domain of 0 < theoretical –log(p) <3 and extrapolating line beyond 3, suggests that the line departs from fitting the empirical –log(*p*) data approximately at 4. Thus, –log(*p*) > 4 is a reasonable empirical threshold for significance for system level analyses.

To minimize confounding due to smoking, the 3,348 CpGs reported as associated with smoking in Christiansen *et al* [101] were removed. This included CpGs replicated in American Indian and African American samples. Another potential confounder on the DNA methylation patterns is the use of immunosuppressive therapies by most patients with SSc. Given the small sample size of this study and the potential risk of overfitting if adjusting for immunosuppressive use, we show instead that the use of immunosuppressive therapies does not heavily bias the association results, although there are several CpGs that show meaningful increases in beta coefficients due to immunosuppressive medication use (Additional file 1: Figs. S6 and S7). The top three CpGs most significantly associated with SSc among immunosuppressive users include cg22196946 in the 5’UTR near *IL15*, cg22187722 in the 5’UTR near *CPVL*, and cg17710334 in an intergenic region. None of these CpGs reaches the empirical threshold for significance among all SSc patients (Additional file 1: Fig. S7).

The power evaluation tool pwrEWAS was used to estimate power as a function of sample size and effect size (Δβ) for two-group comparisons of DNAm assessed using Illumina Human Methylation BeadChip technology [103]. For a total number of 24 subjects (1:1 case:control ratio), and using liver as a proxy for a homogeneous cell population, this study has 4% power to detect differences in up to 6% in CpG-specific methylation across 20 CpGs between groups (the median DNA methylation difference between averaged methylation levels between cases and controls reported in Table 2), and 6% power to detect differences in up to 8% in CpG-specific methylation across 20 CpGs between groups (the maximum DNA methylation difference between averaged methylation levels between cases and controls reported in Table 2). Hence, this study is not powered to detect the DNA methylation differences reported in Table 2.

Associated CpGs identified as significant at FDR-corrected *p* < 0.4 were used to identify the closest gene and then those genes were analyzed for common pathways and functions. Given the limited statistical power, exploratory analysis of clinical features were not computed.

### RNA sequencing

Total RNA was extracted from classical monocytes isolated from 16 female African American patients and 18 female African American controls using the RNeasy Kits (Qiagen, Germantown, MD, USA) according to the manufacturer’s guideline. RNA integrity (RIN) was verified on an Agilent 2200 TapeStation (Agilent Technologies, Palo Alto, CA, USA). The RIN values ranged from 1.8 to 9.0 with an average RIN of 5.8. Due to the low RIN, RNA sequencing libraries were prepared using Illumina’s TruSeq RNA Exome kit. Total RNA (40 ng) of was used to prepare RNA-Seq libraries following the protocol as described by the manufacturer (Illumina, San Diego, CA, USA). Libraries were clustered at a concentration to ensure an average of 25 million reads per sample on the cBot as described by the manufacturer (Illumina, San Diego, CA, USA). Clustered RNA-seq libraries were single read sequenced using version 4 with 1X50 cycles on an Illumina HiSeq2500. Demultiplexing was performed utilizing bcl2fastq-v2.19 to generate Fastq files.

### Gene expression analysis

Upon sequencing, data was analyzed by Rosalind, with a HyperScale architecture developed by OnRamp BioInformatics. Individual sample reads were aligned to the hg19 reference genome using STAR and quantified using HTseq. The subsequent transcript data was imported into R and the edgeR package used to preprocess the data. The transcript counts were normalized using the trimmed mean of M values (TMM) method. To account for potential non-biological variance in the gene expression data, a quasi-likelihood negative binomial generalized log-linear model [104] was used and the data tested for differential expression. FDR *p* values were then calculated for each transcript. Since no differentially expressed transcripts were identified with FDR-corrected *p* < 0.05, an FDR-corrected *p* < 0.4 was used. The rationale for the FDR setting was guided by the desire to perform a system-level analysis and include as many transcripts as possible, as well as previous studies demonstrating that this threshold permits a sensitive analysis at a system level of genes that are relevant to the underlying biology of the trait [40, 41].

RNASeqPower (version 1.39.0) was used to estimate the power given 16 cases and 18 controls, coverage depth (25 M reads) and a coefficient of variation of 0.4 as recommended for human samples and equivalent across groups [105]. Using an α = 0.0001, which based on our experience is a a good approximation to a FDR = 0.05, this study has 9%, 70%, and 97% power to detect differential expression or fold change (FC) of 1.5, 2.0, and 2.5 or higher, respectively. These FC correspond to logFC >|0.18|, logFC >|0.30|, and logFC >|0.40|, respectively. Given the median logFC ∼|0.40|, this study is powered to detect a reasonable fraction of the differentially expressed transcripts between cases and controls.

### Expression quantitative trait methylation (eQTM) analysis

To identify associations between DNA methylation levels and gene expression of nearby genes, a linear regression model was created using the top CpGs and transcripts from both analyses, ranked by unadjusted *p*-value. A total of 1,272 differentially expressed transcripts that met an FDR-corrected *p* < 0.4 were used for this analysis. To correct for unknown sources of variation in the methylation beta scores (e.g., admixture, cellular contamination), the effects of the first two principal components were regressed out and the residuals used in downstream analysis. To correct for the variation in the RNA-seq data, the data was normalized using the TMM. CpGs within 100 kb of the transcript start or end positions were associated with the respective transcript. The RNA transcript was considered the random effect, and the methylation beta value and age of the patient were considered the fixed effects:

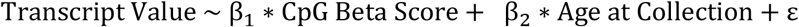

### Functional annotation enrichment analysis

The position of each CpG was annotated to the corresponding genomic location as provided by Illumina (TSS1500, TSS200, 5′ UTR, 1^st^ Exon, Body, Exon boundaries, 3′ UTR, and intergenic). To investigate the distribution of differentially methylated CpGs (DMC) in different genomic locations, the top 100 CpGs were used to compare their localization in different genomic locations. Odds ratio (OR), 95% confidence intervals (CI), and *p*-values were computed against the general distribution of all CpGs of our dataset using GraphPad Prism (version 9.3.1). For regulatory annotation of the differentially methylated CpGs, eFORGE v2.0 (https://eforge.altiusinstitute.org/) [42-44] was used to identify if the associated CpGs were enriched in cell-specific regulatory elements, namely DNase I hypersensitive sites (DHSs) (markers of active regulatory regions) and loci with overlapping histone modifications (H3Kme1, H3Kme4, H3K9me3, H3K27me3, and H3K36me3) across available cell lines and tissues from the Roadmap Epigenomics Project, BLUEPRINT Epigenome, and ENCODE (Encyclopedia of DNA Elements) consortia data. Both the top 100, as well as the top differentially methylated CpGs that met an FDR-adjusted *p* < 0.4, were entered as input of the eFORGE v2.0 analysis and tested for enrichment for overlap with putative functional elements compared to matched background CpGs. The matched background is a set of the same number of CpGs as the test set, matched for gene relationship and CpG island relationship annotation. One thousand matched background sets were applied. The enrichment analysis was completed for different tissues, since functional elements may differ across tissues. Enrichment outside the 99.9th percentile (−log_10_ binomial *p*-value ≥ 3.38) was considered statistically significant (red in Fig. 2).

For Gene Ontology (GO) and functional enrichment analysis, the Database for Annotation, Visualization and Integrated Discovery (DAVID) v6.8 [45, 46], and the Enrichr [47, 48] tools were used. For GO analysis, DAVID v6.8 was used via the web interface (https://david.ncifcrf.gov/) using default settings. The gene lists corresponding to the top differentially methylated CpGs, the top differentially expressed transcripts from the RNAseq analysis, and top differentially expressed transcripts from the eQTM analysis (shown in Tables 2, 3, and 4, respectively) were used. Enrichr was also used via the web interface (https://maayanlab.cloud/Enrichr/), using the default settings and the whole genome set as background. The gene lists in Tables 2, 3, and 4 were used for Biological Process and Pathway enrichment. As output, results were exported from the GO Biological Process 2021 and the KEGG 2021 Pathways databases; p-values are reported.

## Data Availability

The gene expression and DNA methylation data presented in this study are openly available in the NCBI Gene Expression Omnibus (GEO) repository (Accession codes GSE196071 for the study, GSE196007 for the DNA methylation, and GSE196070 for the transcriptomic data). All data are also available from the authors on request. All code to reproduce these analyses is available at https://github.com/PeterCAllen/scleroderma_monocyte_analysis.

## List of abbreviations

CpGs: cytosine-phosphate-guanine sites
DEG: differentially expressed genes
DMC: differentially methylated CpG
eQTM: expression quantitative trait methylation
lcSSc: limited cutaneous SSc
dcSSc: diffuse cutaneous SSc
FDR: false discovery rate
SSc: systemic sclerosis

## Declarations

### Ethics approval and consent to participate

This study was approved by the Institutional Review Board at the Medical University of South Carolina (Pro# 33636). Informed consent was obtained from all participants. All research included in this manuscript conforms with the Declaration of Helsinki.

### Consent for publication

Not applicable.

### Authors’ contributions

Conceptualization: PSR; Formal analysis: PCA, SS, DBF; Software: PCA; Investigation: PCA, SS, RCW, JRW, NHW, JF; Data curation: PCA; Visualization: PCA, SS, DBF; Resources: DMA, GSG, MAC; Supervision: DMA, MAC, CDL, PSR; Writing (original draft): SS, PSR; Writing (review and editing): SS, PCA, CDL, PSR. Project administration: PSR, DMA; Funding acquisition: PSR, GSG. All authors read and approved the final manuscript.

## Acknowledgments

The authors would like to acknowledge all the participants for their contribution to the study.

