## Additional File for "Distinct genome-wide DNA methylation and gene expression signatures in classical monocytes from African American patients with systemic sclerosis"

Additional Information for

**This PDF file includes:**

Figures S1 to S7

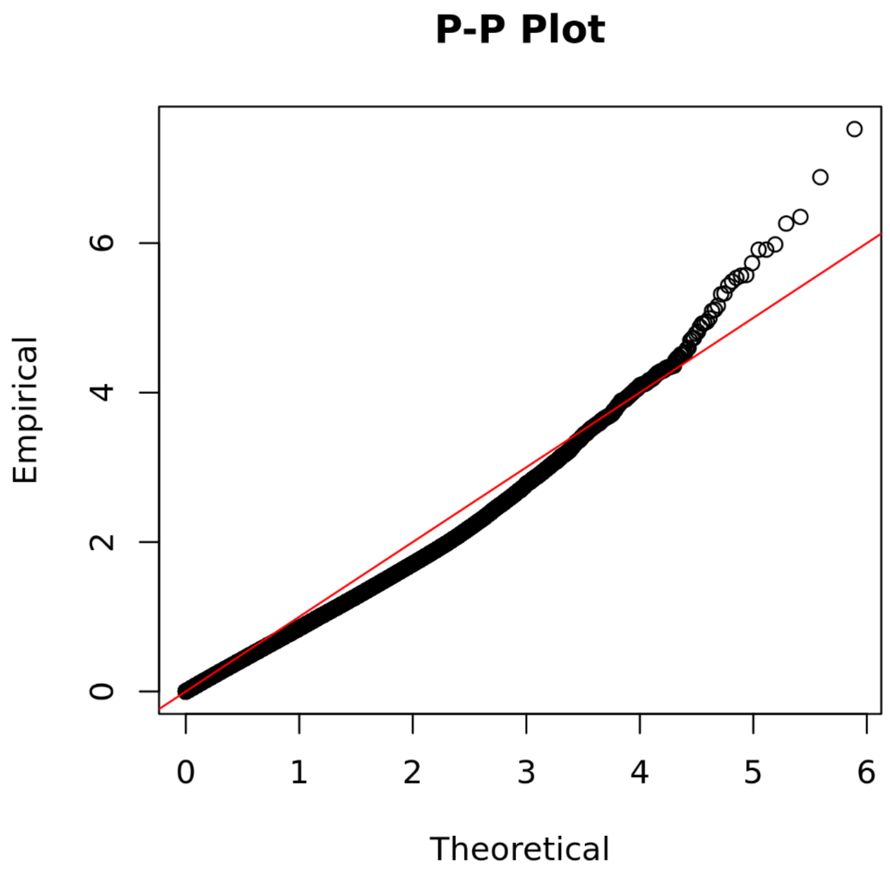


**Figure S1**. P-P plot of CpG association testing results. p values of correlation are plotted for all CpGs as a function of the normal distribution. The red line is equal to the expectation on H0. Fitting a linear regression line for the theoretical domain of 0 < theoretical –log(p) <3 and extrapolating line beyond 3, suggests that the line departs from fitting the empirical –log(p) data approximately at 4, suggesting –log(*p*) > 4 provides a potentially interesting subset for system level analyses such as pathway analysis.

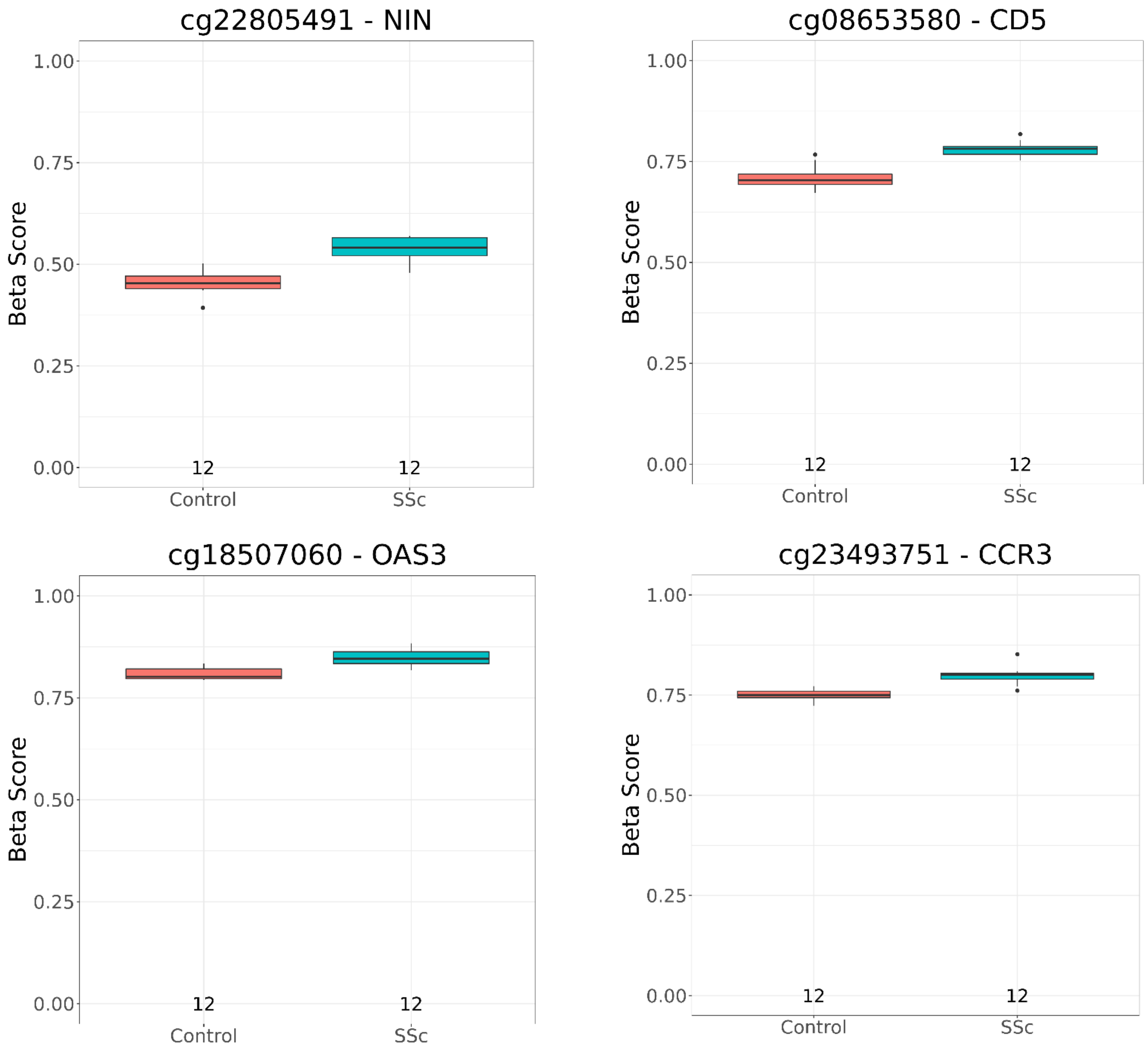


**Figure S2.** Box plots of DNA methylation β-values for cases and controls for four CpG sites selected from Table 2.

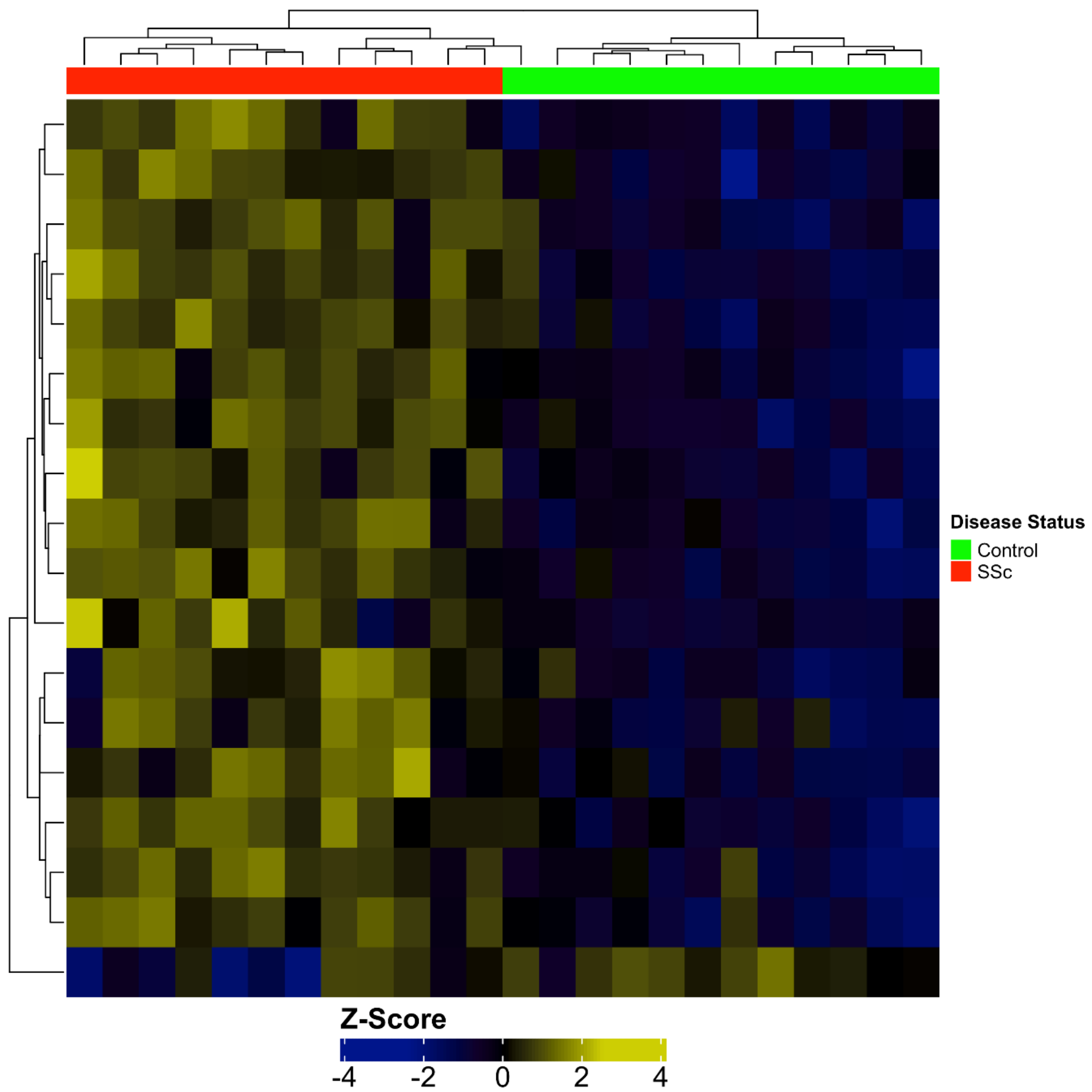


**Figure S3.** Hierarchical cluster analysis (heat map) for the CpGs that passed FDR < 0.4 between cases (red columns) and controls (green columns). The z-score for each CpG is shown, defined as follows: $z = (x - \mu) / \sigma$ where x was the observed value (methylation beta value of CpG for that sample), $\mu$ was the population mean of that CpG, and $\sigma$ was the population standard deviation of the CpG. Blue indicates how many standard deviations below the population mean of that CpG and yellow indicates how many standard deviations above the population mean for that CpG.

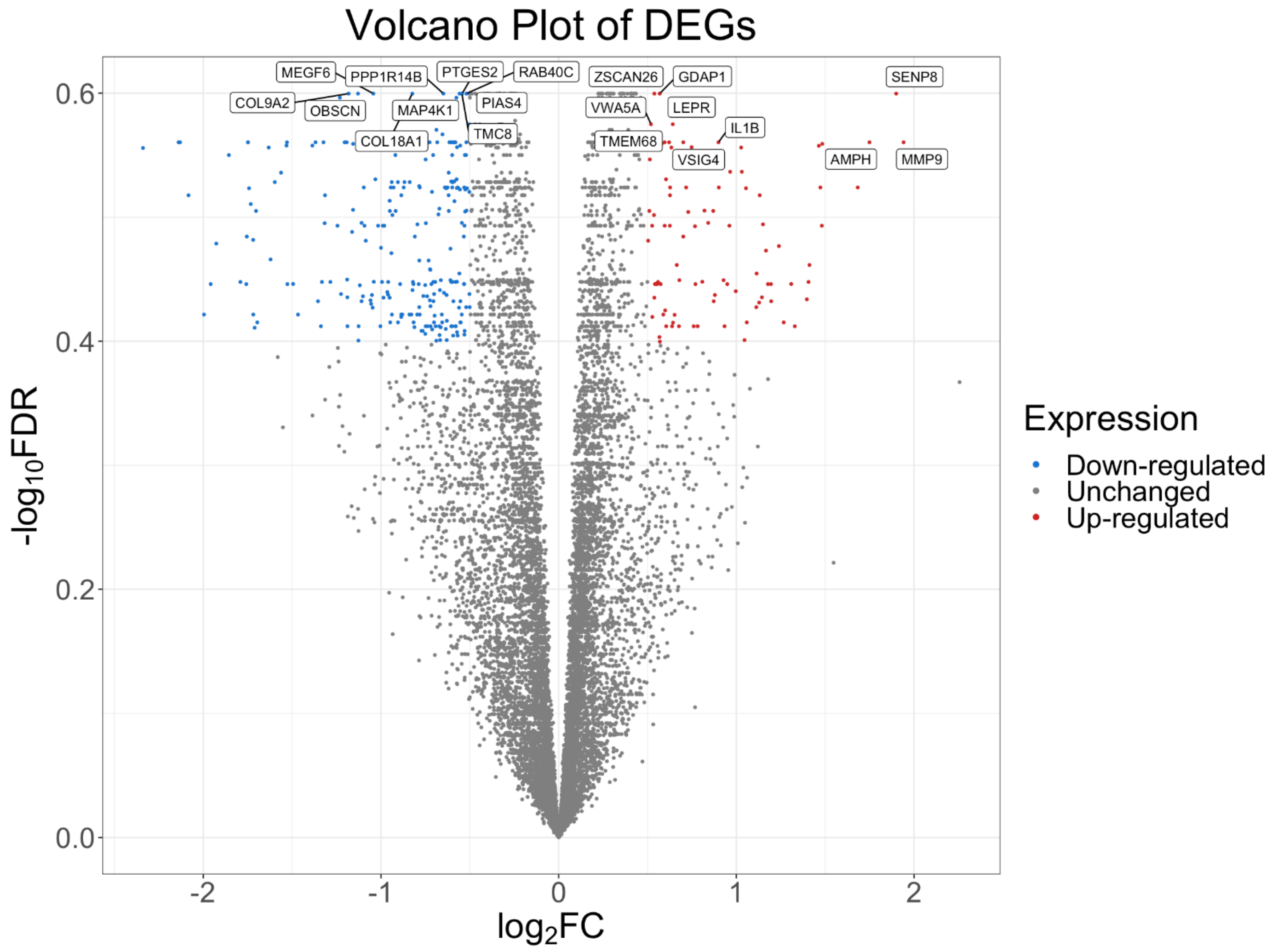


**Figure S4.** Volcano plot displaying differentially expressed genes between African American female patients with scleroderma and controls.

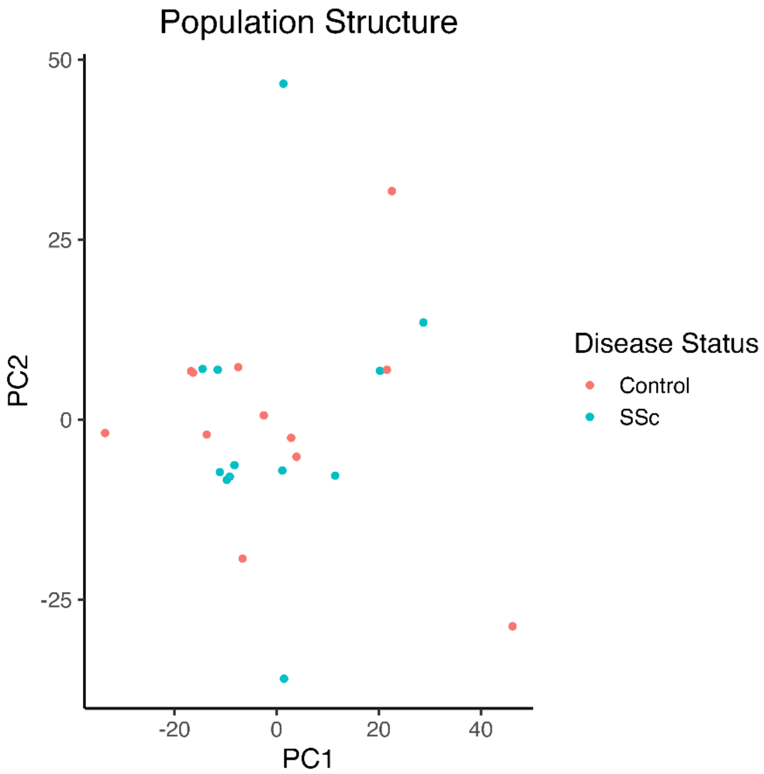


**Figure S5.** First two PCs of the methylation levels of patients with SSc and controls.

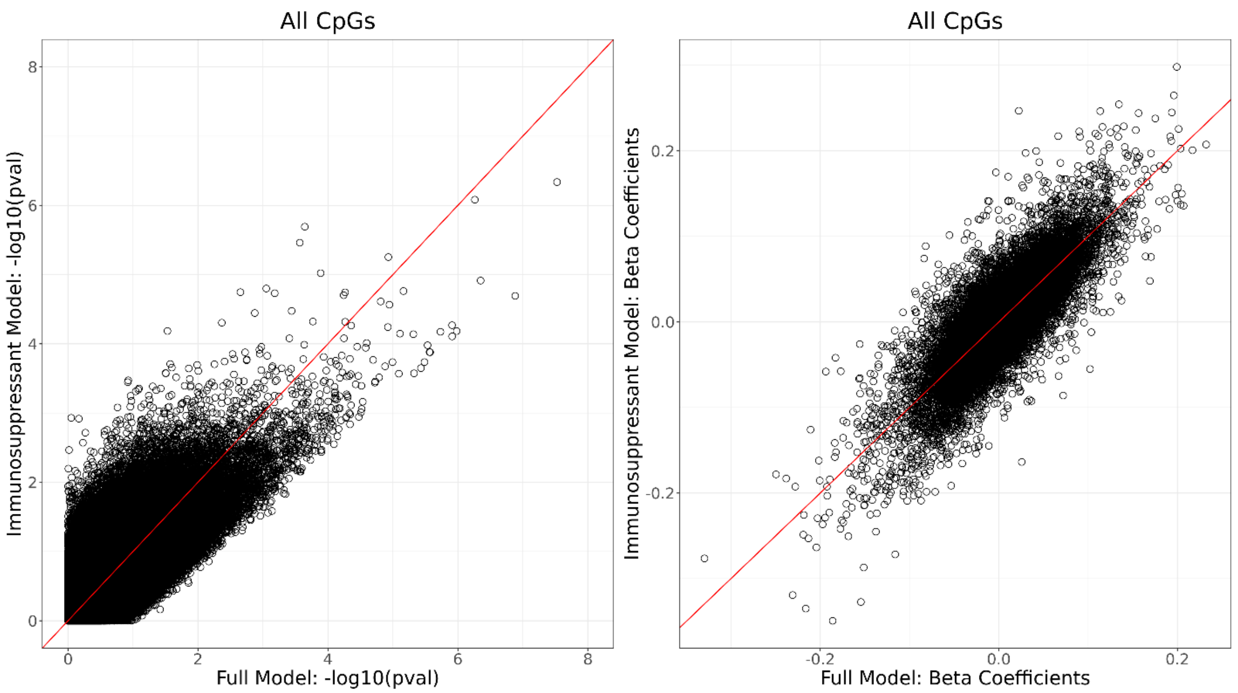


**Figure S6**. Comparison of the analysis of the methylomes of the 8 patients on immunosuppressants and 12 controls (Immunosuppressant Model; y axis) to the analysis of the methylomes of all 12 patients and 12 controls (Full Model; x axis). As shown in the plot of the -log(p) (left side), the results are mostly aligning with the line of unity, not heavily biased by immunosuppressant use. Same for the plot of beta coefficients (right side). These plots show that there aren't substantial global differences in the methylation effects when just looking at the immunosuppressant cases and the combined set of cases.

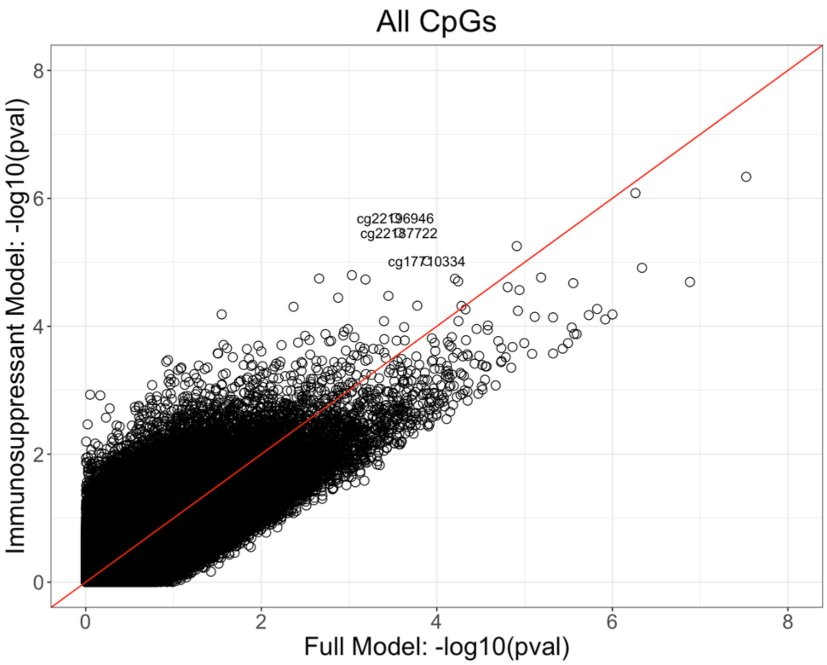


**Figure S7.** The top three CpGs most significantly associated in the Immunosuppressant Model include cg22196946 in the 5'UTR near *IL15*, cg22187722 in the 5'UTR near *CPVL*, and cg17710334 in an intergenic region. None of these CpGs reaches the empirical threshold for significance in the Full Model.
